# Evaluation of the Roche Elecsys Anti-SARS-CoV-2 Assay

**DOI:** 10.1101/2020.06.28.20142232

**Authors:** CS Lau, SP Hoo, SF Yew, SK Ong, LT Lum, PY Heng, JG Tan, MS Wong, TC Aw

## Abstract

**Background:** Little is known about the performance of the Roche novel severe acute respiratory syndrome coronavirus 2 antibody (anti-SARS-CoV-2) assay. We provide an extensive evaluation of this fully automated assay on the Cobas e801/e602 immunoassay analysers.

**Methods:** We assessed the linearity, precision, and throughput of the Roche anti-SARS-CoV-2 assay. Sensitivity was calculated from 349 SARS-CoV-2 polymerase chain reaction (PCR) positive samples; specificity was determined from 714 coronavirus disease 2019 (COVID-19)-naive samples. We examined cross-reactivity against other antibody positive samples (syphilis, RF, ANA, ds-DNA, influenza, dengue, HBV, HCV) and the anti-SARS-CoV-2 kinetics.

**Results:** The assay cut-off index (COI) was linear up to 90.7. The inter-assay precision was 2.9% for a negative control (COI=0.1) and 5.1% for a positive control (COI=3.0). Assay time is 18min and results are available 1 minute later; throughput for 300 samples was 76 minutes. No cross-reactivity was observed with other antibody positive samples; specificity was 100%. The assay has a sensitivity of 97.1% 14 days after PCR positivity (POS) and 100% at ≥21 days POS; 48.2% of cases had anti-SARS-CoV-2 within 6 days POS. In 11 subjects in whom serum was available prior to a positive antibody signal (COI ≥1.0) the interval between the last negative and first positive COI (time to “sero-conversion”) on average is 3 days (range 1-6 days) and 4 more days (range 1-7) for the anti-SARS-CoV-2 to plateau.

**Conclusion:** The Roche anti-SARS-CoV-2 assay shows excellent performance with minimal cross-reactivity from other viral and confounding antibodies. Antibody development and sero-conversion appears quite early.

## INTRODUCTION

The novel severe acute respiratory syndrome coronavirus 2 (SARS-CoV-2) has caused a global pandemic. Coronavirus disease 2019 (COVID-19) can present with a range of symptoms (*1, 2*) and inflicts a disturbingly higher fatality rate in older patients (*3*). Viral testing (nucleic acid) remains the recommended diagnostic test for infection (*4, 5*). However, its long turn-around time has led to calls for more rapid tests like serology. Antibody testing can act as an indirect marker for infection, help identify patients with prior infection or exposure and can be performed relatively quickly. An immune response can develop (*6*) as soon as 10-13 days after a COVID-19 infection (*7, 8*), However, the early rapid point-of-care tests (POCT) for anti-SARS-CoV-2 were fraught with inaccuracy. Roche Diagnostics has developed a SARS-CoV-2 antibody (anti-SARS-CoV-2) assay for use on its electrochemiluminescent-immunoassay analysers for the rapid, continuous testing of large numbers of samples. This assay has received approval from the Food and Drug Administration (FDA) (*9*) for laboratory use in May 2020. Some brief evaluations of this assay have been recently available as editorial letters (*10, 11*). We describe a more extensive evaluation of the Roche anti-SARS-CoV-2 assay on Cobas immunoassay analyzers across 3 institutions in Singapore.

## MATERIALS AND METHODS

### Patient cohorts

Residual serum samples from cases with suspected or confirmed SARS-CoV-2 infection from April to June 2020 were recruited from 3 institutions in Singapore: Changi General Hospital, Khoo Teck Puat Hospital and Sengkang General Hospital. For evaluating diagnostic sensitivity, 415 excess serum samples from COVID-19 cases (from 280 individual patients) testing positive for SARS-CoV-2 by PCR were used. For sensitivity analysis of an evolving biomarker like anti-SARS-CoV-2 at different time points in the disease evolution, it is customary to divide subjects by days from onset of symptoms or by days post initial PCR positivity (POS). We elected to use POS rather than onset of symptoms as it is a more objective milestone and precludes confounding issues with patients who maybe asymptomatic or pre-symptomatic. Antibody positive samples [dengue N=74, hepatitis C (HCV) N=3, hepatitis B (HBV) N=12, syphilis N=1, antinuclear antibody (ANA) N=16, double-stranded DNA antibody (anti-ds-DNA) N=4, rheumatoid factor (RF) N=7] were recruited for cross-reactivity analysis; these samples were from ambulatory subjects with no suspicion for COVID-19 or acute respiratory illness. For diagnostic specificity, samples were obtained from 597 consenting healthcare workers (HCWs), 315 of whom had received their annual southern hemisphere influenza vaccination 4 weeks prior to testing. All samples were collected in plain serum tubes.

As the Roche anti-SARS-CoV-2 assay is a qualitative test, its sensitivity can be represented by the positive percentage agreement (PPA) between antibody positivity against all PCR positive patients tested. Conversely, the specificity of the test is represented by the negative percentage agreement (NPA) between antibody negativity against all control subjects.

To study sero-conversion, we examined SARS-CoV-2 PCR positive subjects with residual sera who were initially non-reactive for anti-SARS-CoV-2 but became reactive later.

### Instrumentation and analysis

The Roche anti-SARS-CoV-2 assay is a sandwich immunoassay where biotinylated SARS-CoV-2 specific recombinant antigens and SARS-CoV-2 specific recombinant antigens labelled with ruthenium form a sandwich complex with anti-SARS-CoV-2. Streptavidin-coated microparticles are then added, and the complex becomes bound to a solid phase via interactions between biotin and streptavidin. Once the microparticles are captured onto the electrode surface and unbound substances are washed away, an electrical current at the electrode induces chemiluminescence. When compared to the mean chemiluminescent signal of a calibrator, an anti-SARS-CoV-2 index is derived with a reported cut-off index (COI) of 1.0 for positivity. Initially the assay did not include any quality control (QC) materials and the manufacturer advised laboratories to prepare their own QC materials. A negative control derived from serum samples with a target COI of <0.8 and a positive control derived from serum samples with a target COI of 3-15 was recommended. Laboratories were also encouraged to watch their internal QCs carefully as the preliminary on-board stability of the reagents was 72 hours. The stated assay specificity in the package insert is 99.8% with a sensitivity of 100% for samples ≥14 days post symptom onset.

The molecular laboratories of all 3 hospitals employed real-time polymerase chain reaction (PCR) test systems that targeted at least 2 viral epitopes of SARS-CoV-2. Changi and Sengkang hospitals employed the Roche Cobas e801 while Khoo Teck Puat used the Cobas e602 immunoassay analyzer. Results from all three hospitals were pooled for sensitivity and specificity analyses while Changi General provided details of the technical analyses (precision, linearity and throughput). Inter-assay precision (CV) was analysed using 5 serum pools (including negative and positive controls prepared as per manufacturer recommendations, and 3 positive serum samples over a range of reactive COI values) run 5 times daily over 5 days, as per the CLSI EP15-A3 protocol (*12*). Assay linearity was assessed on several pools of reactive samples covering the clinically relevant range of COI. The laboratories at Changi General and Khoo Teck Puat are accredited by the College of American Pathologists.

Statistical analyses were performed using MedCalc software v19.3.1 (MedCalc, Ostend, Belgium). As this work was part of routine evaluation of new diagnostic assays, it was deemed exempt by our institutional review board.

## RESULTS

### Precision and linearity

The Roche assay showed good precision, with a CV of 2.9% for negative controls (mean COI = 0.1) and 5.1% for positive controls (mean COI = 3.0) (See Table 1); at higher COI values (9.1, 18.9 and 42.4), the precision was very good (1.4%, 1.5%, 2.0%). The assay COI signal was found to be linear from 1.0 to 90.7 (y = 181.44x – 1.3568) and curvilinear rightwards thereafter (see Figure 1, Supplementary Table 1).

**Table 1:** Precision of the Roche anti-SARS-CoV-2 assay

| Pool ID | Pool type | Min Index | Max Index | Mean COI | SD | %CV |
| --- | --- | --- | --- | --- | --- | --- |
| 1 | Negative control (patient sera) | 0.074 | 0.084 | 0.078 | 0.002 | 2.87 |
| 2 | Positive control (patient sera) | 2.770 | 3.280 | 2.991 | 0.153 | 5.11 |
| 3 | S1 | 8.840 | 9.260 | 9.065 | 0.129 | 1.43 |
| 4 | S2 | 18.200 | 19.400 | 18.856 | 0.277 | 1.47 |
| 5 | S3 | 40.700 | 43.700 | 42.444 | 0.850 | 2.00 |

**Figure 1:**
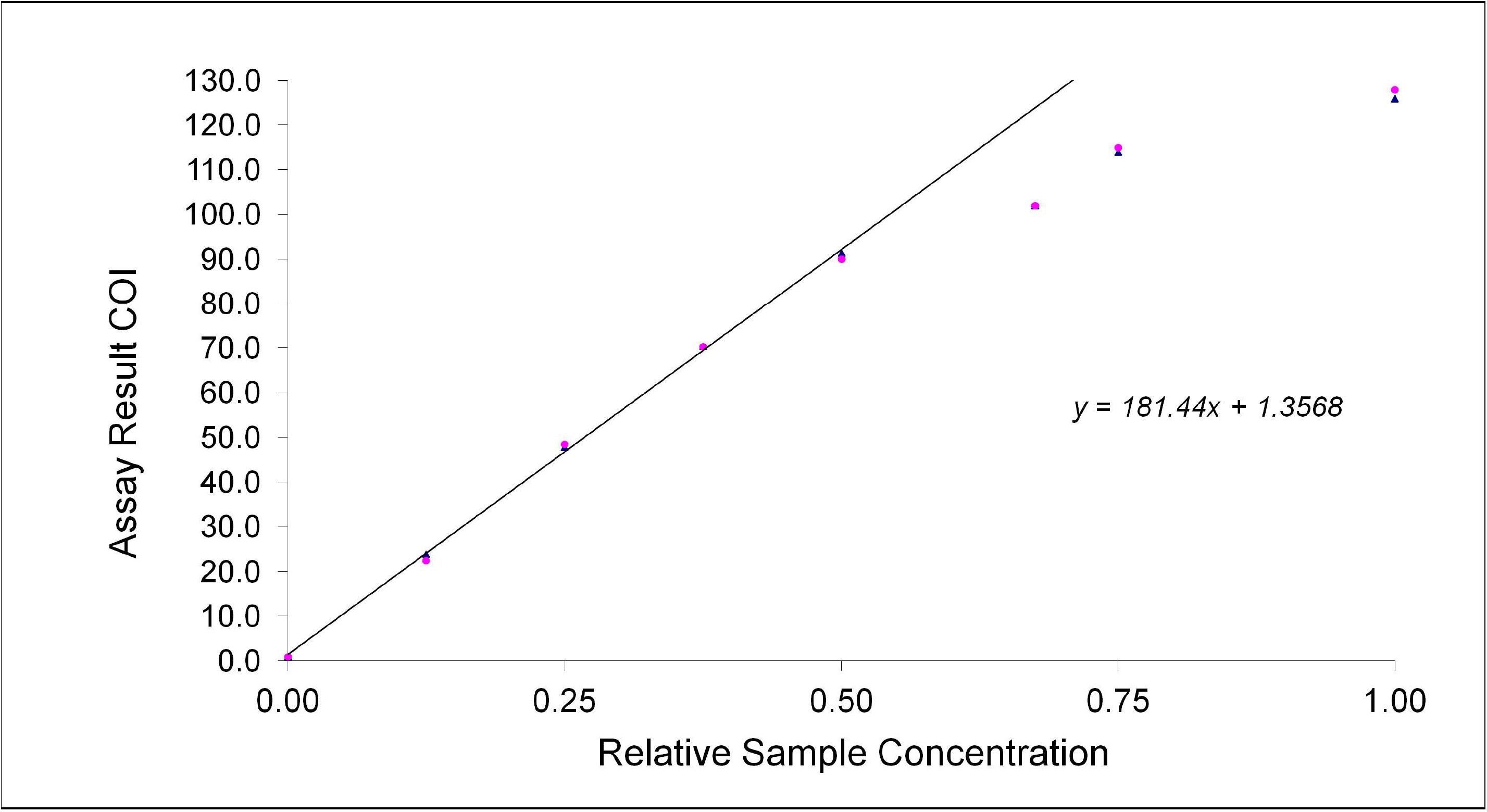
Linearity plot of the Roche anti-SARS-CoV-2 assay.

### Throughput

At Changi General the anti-SARS-CoV-2 assay on the Roche Cobas e801 is run using two measuring cells. The Cobas e801 was able to analyse 50 samples in 29 minutes and 100 specimens in 39 minutes. When loaded with 300 samples (and not performing other routine tests simultaneously), the Cobas e801 completed the sample analysis in 76 minutes (see Supplementary Table 2).

**Table 2:**
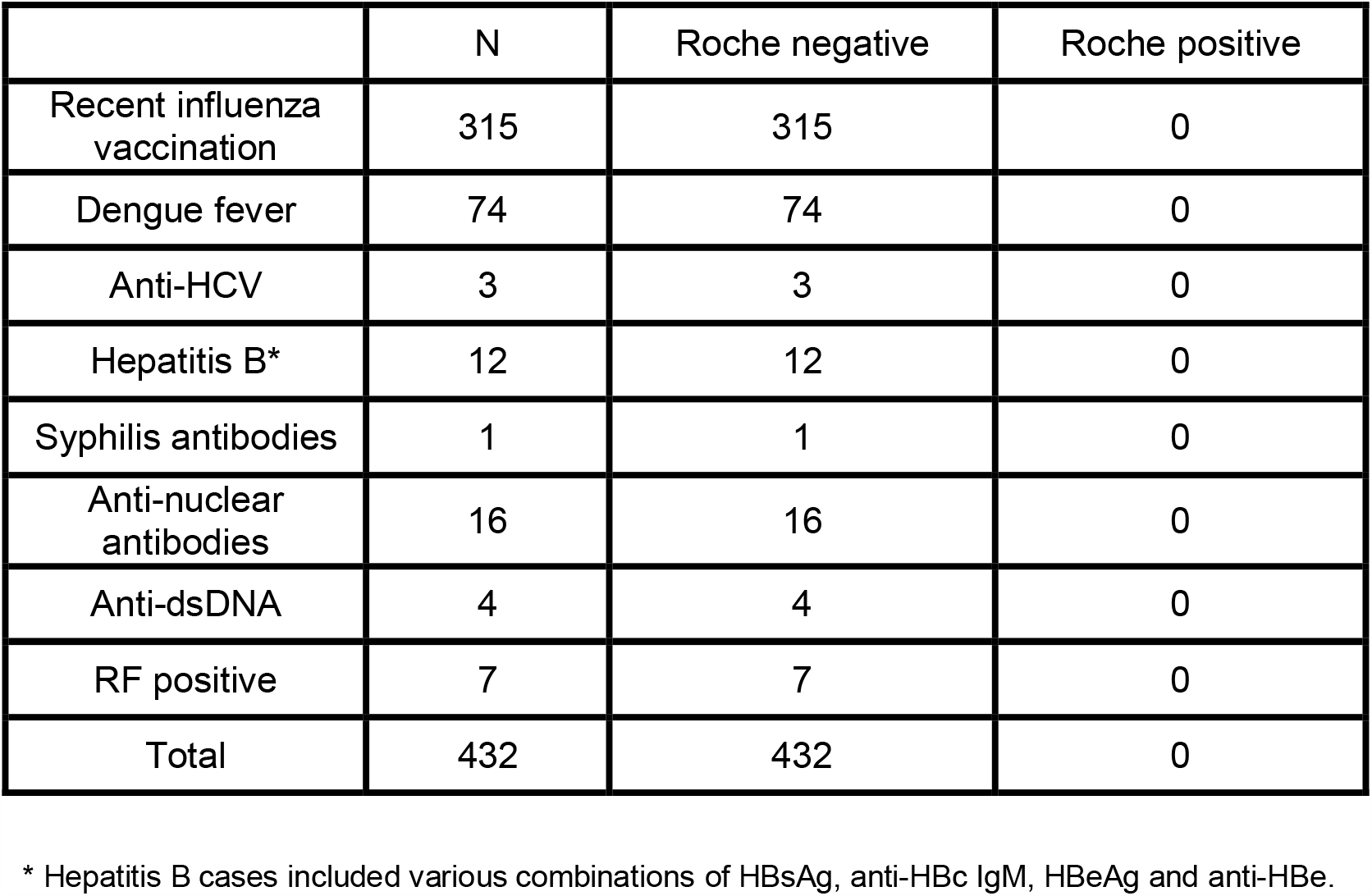
Cross-reactivity analysis for the Roche anti-SARS-CoV-2 assay.

### Cross-reactivity

Out of the 315 HCWs who received their annual influenza vaccination 4 weeks prior to testing, none were reactive for anti-SARS-CoV-2. All cases positive for other antibodies (n=117) also tested negative for anti-SARS-CoV-2 (see Table 2).

### Specificity

As there was no cross-reactivity with anti-SARS-CoV-2 in the 432 samples above (Table 2), we included them in diagnostic specificity analysis together with another 282 samples from other HCWs. None of these 714 samples were reactive on the Roche anti-SARS-CoV-2 assay, yielding a specificity of 100.00% (95% CI 99.47 to 100.00).

### Sensitivity

Out of the samples recruited for diagnostic sensitivity testing (N=415), 66 were residual samples from inpatients (when they were not initially suspected of having COVID-19) but who subsequently tested positive for SARS-CoV-2 PCR. Therefore, these 66 samples were excluded from the sensitivity analysis. Of the remaining 349 samples (from 205 individual patients) the PPA increased to 97.1% after 14 days post positive PCR (POS), and to 100% after 21 days POS (see Table 3). Notably, 48.2% of patients had positive antibodies within the first week after their PCR diagnosis. Interestingly, of the 66 samples above (excluded from the sensitivity analyses) with late PCR diagnosis 48.5% were positive for anti-SARS-CoV-2.

**Table 3:**
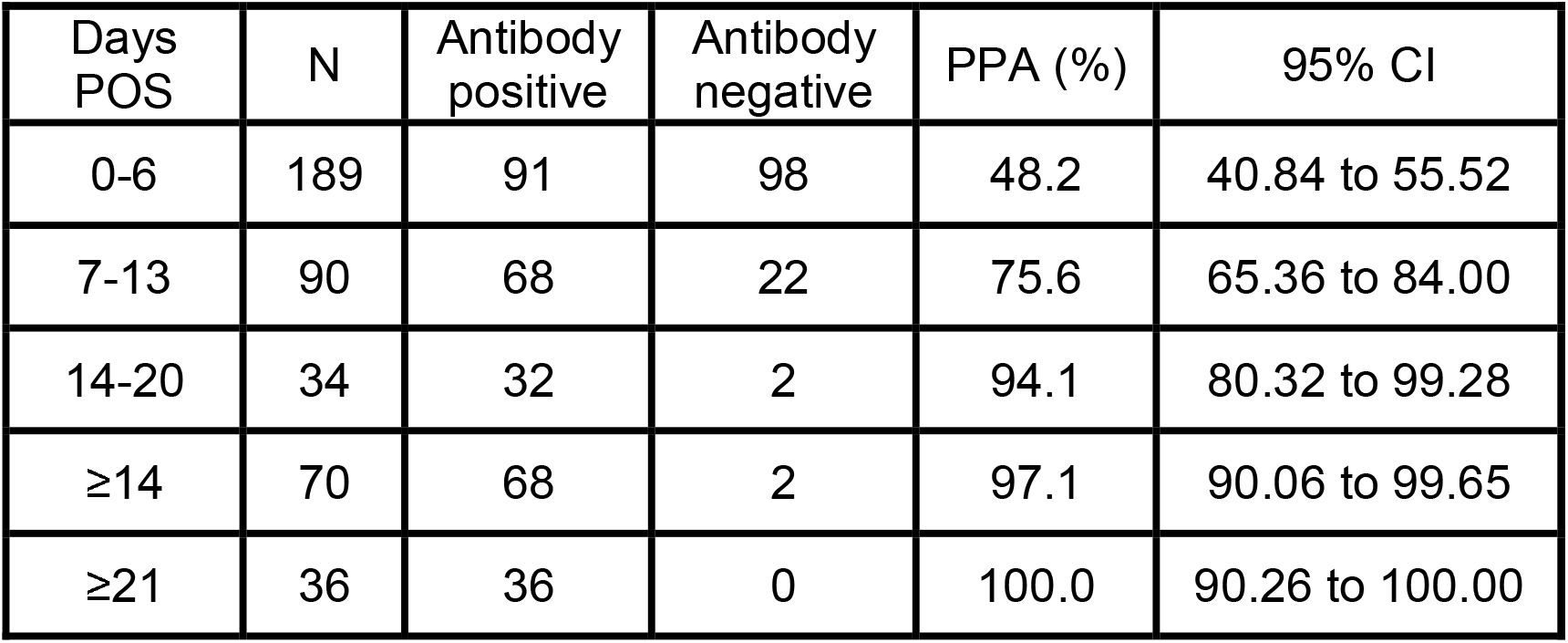
Roche assay sensitivity (positive percentage agreement) by days post positive PCR.

### Positive and negative predictive values

Assuming a disease prevalence of 5%, the predictive value of the Roche anti-SARS-CoV-2 assay (specificity of 100% and sensitivity of >97% when used after 14 days POS) will be as follows: positive predictive value (PPV) 100.00% and negative predictive value (NPV) 99.9% (95%CI 99.41, 99.96); MedCalc did not provide the CI for the PPV.

### Sero-conversion

Eleven subjects were available for sero-conversion study and the progression of their antibody levels (COI) were plotted against days POS (See Figure 2 and Table 4). Using the manufacturer recommended COI of 1.0 the interval for “sero-conversion” (last negative reading to first positive reading), extrapolated from figure 2, ranged from 1-6 days with an average of 3 days. In 8 of the 11 subjects the antibody levels plateaued over time (1-7 days from positivity, mean of 4 days).

**Table 4:** SARS-CoV-2-Ab COI trends by days POS in patients with initial COI <1.0.

| Patient | Days POS | COI | POS day when COI $\geq 1.0^*$ | Interval between negative to COI $\geq 1.0$ | POS day of plateau* | Interval to plateau* |
| --- | --- | --- | --- | --- | --- | --- |
| A | 0 | 0.08 | 4+ | 5 | 11+ | 7 |
|  | 2 | 0.09 |  |  |  |  |
|  | 7 | 7.59 |  |  |  |  |
|  | 11 | 9.98 |  |  |  |  |
| B | 0 | 0.08 | 2+ | 3 | NA | NA |
|  | 1 | 0.09 |  |  |  |  |
|  | 4 | 6.03 |  |  |  |  |
|  | 6 | 21.0 |  |  |  |  |
| C | 1 | 0.09 | 3+ | 4 | 5+ | 2 |
|  | 5 | 5.44 |  |  |  |  |
|  | 9 | 6.59 |  |  |  |  |
|  | 10 | 5.03 |  |  |  |  |
| D | 3 | 0.08 | 9+ | 2 | 12+ | 3 |
|  | 7 | 0.09 |  |  |  |  |
|  | 8 | 0.12 |  |  |  |  |
|  | 10 | 2.49 |  |  |  |  |
|  | 12 | 13.8 |  |  |  |  |
|  | 14 | 5.86 |  |  |  |  |
|  | 17 | 7.19 |  |  |  |  |
|  | 19 | 12.2 |  |  |  |  |
| E | 0 | 0.08 | 5+ | 2 | 11+ | 6 |
|  | 1 | 0.09 |  |  |  |  |
|  | 5 | 0.51 |  |  |  |  |
|  | 7 | 2.36 |  |  |  |  |
|  | 11 | 12.6 |  |  |  |  |
|  | 15 | 13.1 |  |  |  |  |
| F | 4 | 0.08 | 6+ | 1 | 10+ | 4 |
|  | 6 | 0.15 |  |  |  |  |
|  | 7 | 1.17 |  |  |  |  |
|  | 10 | 14.7 |  |  |  |  |
|  | 14 | 15.9 |  |  |  |  |
|  | 16 | 14.3 |  |  |  |  |
| G | 2 | 0.08 | 15+ | 6 | NA | NA |
|  | 6 | 0.19 |  |  |  |  |
|  | 12 | 0.35 |  |  |  |  |
|  | 18 | 2.72 |  |  |  |  |
| H | 0 | 0.07 | 5+ | 2 | 9+ | 4 |
|  | 1 | 0.07 |  |  |  |  |
|  | 2 | 0.07 |  |  |  |  |
|  | 3 | 0.11 |  |  |  |  |
|  | 4 | 0.13 |  |  |  |  |
|  | 6 | 6.26 |  |  |  |  |
|  | 7 | 9.27 |  |  |  |  |
|  | 8 | 13.9 |  |  |  |  |
|  | 9 | 17.8 |  |  |  |  |
|  | 12 | 18.6 |  |  |  |  |
|  | 15 | 12.7 |  |  |  |  |
|  | 19 | 10.4 |  |  |  |  |
| I | 0 | 0.11 | 2+ | 4 | NA | NA |
|  | 1 | 0.19 |  |  |  |  |
|  | 5 | 7.98 |  |  |  |  |
| J | 2 | 0.12 | 4+ | 2 | 8+ | 4 |
|  | 4 | 0.34 |  |  |  |  |
|  | 6 | 10.3 |  |  |  |  |
|  | 8 | 27.6 |  |  |  |  |
|  | 10 | 26.9 |  |  |  |  |
|  | 13 | 38.5 |  |  |  |  |
| K | 2 | 0.12 | 3+ | 2 | 4+ | 1 |
|  | 4 | 1.99 |  |  |  |  |
|  | 9 | 2.11 |  |  |  |  |
|  | 12 | 1.76 |  |  |  |  |
|  | 14 | 1.59 |  |  |  |  |
|  | 18 | 1.96 |  |  |  |  |
|  | 22 | 4.24 |  |  |  |  |

\* based on interpolation from figure 2.

**Figure 2:**
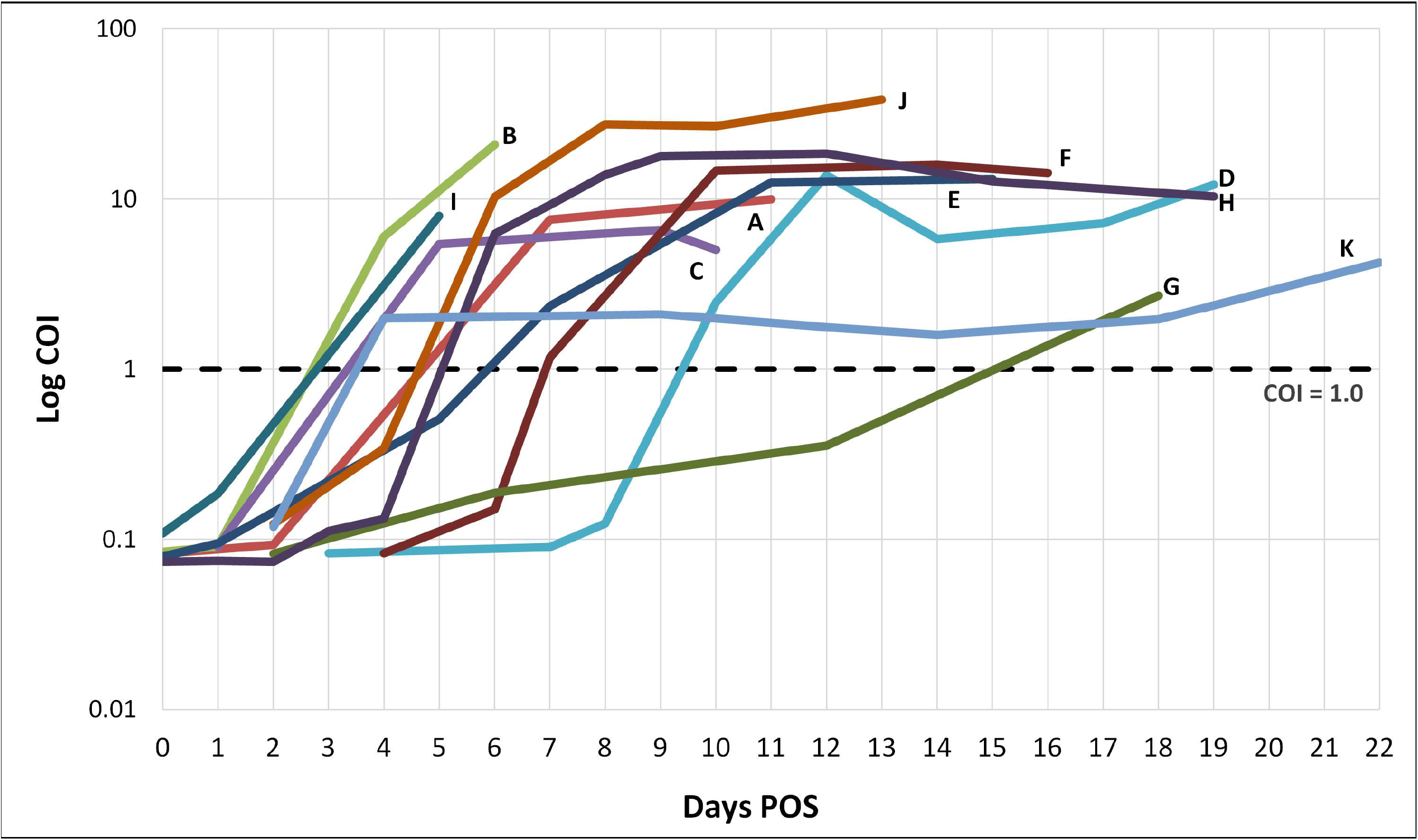
SARS-CoV-2-Ab COI kinetics by days POS in patients with initial COI <1.0.

## DISCUSSION

The Roche anti-SARS-CoV-2 assay shows good precision (CV of 2.9% and 5.1% at COI levels of 0.1 and 3.0) and a linear signal response over a wide COI range (0.98 to 90.7). The diagnostic sensitivity of the assay is 97.1% ≥14 days POS, or 100% after 21 days. The diagnostic specificity is 100.0% in 714 non-COVID-19 samples tested. The PPV was 100.00%, and NPV 99.9% at ≥14 days POS. Our results are in close agreement with the manufacturer’s reported specifications. They are also in agreement with other evaluations of the Roche anti-SARS-CoV-2 assay in the literature (*10, 11, 13*); the reported assay CV (2.1-4.6%, 1.2-3.6%, <3.9%), test sensitivity at ≥14 days post-onset of symptoms (87.7%, 91.1%, 89.4%) and test specificity (100%, 100%, 98.7%) were similar to ours. This assay easily meets the US Centers for Disease Control and Prevention (CDC) and the FDA requirements for an acceptable SARS-CoV-2 serology test - >90% sensitivity and >95% specificity (*14, 15*). As the sensitivity/PPA only increases to over 90% after 14 days post PCR positivity, patients should not be tested earlier. This is also supported by other studies showing that it takes up to 11-14 days before anti-SARS-CoV-2 develop (*16*). Furthermore, the Roche assay has an excellent throughput, analysing 300 samples in 76 minutes. This is a major boon when processing large sample numbers from screening centres.

In our study we have used days POS to categorize our population. As COVID-19 can have asymptomatic and pre-symptomatic patients (*17-19*), using the presence of symptoms as a pre-selection criterion for inclusion will exclude asymptomatic and pre-symptomatic cases; days POS will mitigate against this. Notably, out of 66 leftover serum samples that were available before COVID-19 was suspected and PCR tested positive, 32 were already positive for anti-SARS-CoV-2. From the 11 patients with multiple longitudinal samples in figure 2, “sero-conversion” (interval between last negative point and first positive point) takes an average of 3 days, and that antibody titres plateau an average of 4 days later. A recent report *(8)* found that the time course of COVID19 antibodies (both IgG and IgM) tended to plateau in 6 days after they become positive in 19 subjects. Anti-SARS-CoV-2 has also been detected on the first day of PCR positivity (*20*) or within 7 days of symptom onset in 12.2% of patients (*8*). This indicates that in some patients, anti-SARS-CoV-2 development can occur earlier than expected.

A recent report (*21*) found that SARS-CoV-2 infection may cause false-positive dengue antibody results. It is noteworthy that in patients with positive dengue serology (n = 74) we did not encounter any false positive anti-SARS-CoV-2. Our study also confirms that the assay remains specific even in patients positive for HCV, HBV, syphilis, ANA, ds-DNA and RF. Moreover, in 315 samples with post-influenza vaccination, none were positive for anti-SARS-CoV-2, reinforcing the fact that the Roche assay has little cross-reactivity with influenza antibodies. The sero-negativity of most of our HCWs also underscores the adequacy of our personal protective equipment (PPE) measures. The PPE measures taken by hospital staff across institutions in Singapore are similar (*22*), and include the use of surgical masks in low-risk areas (non-COVID-19 inpatient rooms, afebrile patient zones in the emergency department, non-infectious disease clinics) and N95 respirators/eye protection/gowns/gloves in high-risk areas (COVID-19 isolation rooms, febrile patient zones in the emergency department). However, their lack of antibodies also means that most of these HCWs remain at risk for future COVID-19 infections and they should continue to rigorously enforce safety precautions.

The strength of this evaluation is that it is a multi-center collaborative effort with sufficient sample numbers for sensitivity, specificity and cross-reactivity analyses. A limitation of our study is that we have relatively fewer samples for cases after 14 days POS. Further studies on larger populations would be desirable. In addition, we do not have the sero-prevalence of SARS-CoV-2 in our community. At the time of this study, we created our own negative and positive control materials from patient sera based on manufacturer recommendations. However, control materials have since been supplied by the manufacturer. The on-board reagent stability of 72 hours in the initial manufacturer information sheet has also been updated to 14 days.

## CONCLUSION

Our results show that the Roche anti-SARS-CoV-2 assay has excellent performance and is highly comparable both with the manufacturer’s information and other published studies. With an excellent throughput, the Cobas e801 should be extremely useful in the tracking the spread of SARS-CoV-2 in various populations and for sero-prevalence studies. We have also found that anti-SARS-CoV-2 can develop early.

## Data Availability

NA

## ABBREVIATIONS

SARS-CoV-2: Novel severe acute respiratory syndrome coronavirus 2
COVID-19: Coronavirus disease 2019
Anti-SARS-CoV-2: SARS-CoV-2 antibody
FDA: Food and Drug Administration
HCV: Hepatitis C
HBV: Hepatitis B
RF: Rheumatoid factor
HCWs: Health care workers
COI: Cut-off index
PCR: Polymerase chain reaction
CV: Inter-assay precision
PPA: Positive percentage agreement
NPA: Negative percentage agreement
POS: Post positive PCR
PPV: Positive predictive value
NPV: Negative predictive value
CDC: Centers for Disease Control and Prevention
PPE: Personal protective equipment
ANA: anti-nuclear antibody
Anti-ds-DNA: double-stranded DNA antibody

